# Right amygdala lesions are associated with improved mood after epilepsy surgery

**DOI:** 10.1101/2022.11.21.22282596

**Authors:** Fatimah M. Albazron, Daniel Tranel, Matthew A. Howard, Aaron D. Boes

**Affiliations:** Department of Pediatrics, Carver College of Medicine, University of Iowa, 200 Hawkins Drive, Iowa City, IA, 52242, USA; Department of Neurology, Carver College of Medicine, University of Iowa, 200 Hawkins Drive, Iowa City, IA, 52242, USA; Department of Psychological and Brain Sciences, University of Iowa, 200 Hawkins Drive, Iowa City, IA, 52242, USA; Department of Neurosurgery, Carver College of Medicine, University of Iowa, 200 Hawkins Drive, Iowa City, IA, 52242, USA; Department of Psychiatry, Carver College of Medicine, University of Iowa, 200 Hawkins Drive, Iowa City, IA, 52242, USA; Iowa Neuroscience Institute, University of Iowa, 200 Hawkins Drive, Iowa City, IA, 52242, USA

**Keywords:** Temporal lobe epilepsy, Lesion symptom mapping, Mood changes, Amygdala

## Abstract

Neuroimaging studies in healthy and clinical populations strongly associate the amygdala with emotion, especially negative emotions. The consequences of surgical lesions of the amygdala on mood are not well characterized. We tested the hypothesis that amygdala lesions would result in mood improvement. In this study we evaluated a cohort of 52 individuals with medial temporal lobectomy for intractable epilepsy who had resections variably involving the amygdala. All individuals achieved good post-surgical seizure control and had pre- and post-surgery mood assessment with the Beck Depression Inventory (BDI) ratings. We manually segmented the surgical resection cavities and performed multivariate lesion-symptom mapping of change in BDI. Our results showed a significant improvement in average mood ratings from pre- to post-surgery across all patients. In partial support of our hypothesis, resection of the right amygdala was significantly associated with mood improvement (r = 0.5, p = 0.008). The lesion-symptom map also showed that resection of the right hippocampus and parahippocampal gyrus was associated with worsened post-surgical mood. Future studies could evaluate this finding prospectively in larger samples while including other neuropsychological outcome measures.

## Introduction

The amygdala is a small almond-shaped structure located in the medial temporal lobe, anterior to the hippocampus. It has been strongly implicated in mood and psychopathology, particularly in negative emotions like fear and anxiety (Phelps and LeDoux, 2005; Price and Drevets, 2009). The amygdala is also implicated in major depression, with prior studies demonstrating that the severity of depressive episodes is associated with larger amygdala volume, increased and prolonged amygdala activity, or an increase in amygdala blood flow (Drevets, 2000, 2001; Siegle *et al*., 2002; Zetzsche *et al*., 2006). In contrast, reduced activity of the amygdala has been associated with improvements in depression or increases in positive mood in healthy individuals (Fu *et al*., 2004; Kraehenmann *et al*., 2015).

Most of these findings linking the amygdala to depression are correlative in nature. Methods that allow stronger causal inferences on the role of the amygdala in mood include electrical brain stimulation and changes that occur in the setting of acquired brain lesions. Scangos and colleagues recently demonstrated that gamma power within the amygdala correlated with depression severity, and the reduction of gamma power by electrical stimulation corresponded with a robust mood improvement (Scangos *et al*., 2021), While there are historical data on the effects of amygdala removal in psychosurgery, much of this work was focused on aggression (Mpakopoulou *et al*., 2008; Zhang *et al*., 2017; Gouveia *et al*., 2021), and the effects on mood are not well characterized.

In the current study, we hypothesized that surgical resection of the amygdala would be associated with an improvement in mood. To test this hypothesis, we evaluated mood outcomes in a clinical sample of patients with epilepsy who underwent medial temporal lobectomy as a treatment for medically refractory epilepsy. The Beck Depression Inventory (BDI) is a self-reported scale that assesses symptoms of depression according to the DSM-IV, and was used as our assessment of mood pre- and post-surgery (change in BDI, ΔBDI). Improvement in mood following surgery was evaluated in relation to the brain regions that were resected using multivariate lesion symptom mapping (Pustina *et al*., 2018).

## Materials and methods

### Participants

Participants were obtained from the Patient Registry of the Division of Behavioral Neurology and Cognitive Neuroscience at the University of Iowa. The cohort was selected from a pool of 61 patients with medically refractory epilepsy who underwent unilateral medial temporal lobectomy between 1983 and 2019, completed pre- and post-surgical BDI assessment in the chronic epoch (≥ 3 months), and completed a post-surgery research-quality structural MRI scan ≥ 2 months after the surgery. We included participants who had a successful surgery with regard to seizure outcome by achieving complete or near complete seizure freedom in the first post-surgical year, with near complete seizure freedom defined as only having rare breakthrough seizures secondary to missed medications or in the presence of other provocations. The cohort included 52 participants after excluding patients who continued to have seizures following surgery (n = 7) or with unknown seizure outcome information (n = 2). 23 participants had a right temporal lobectomy (44%). The demographics of all participants are presented in table 1.

**Table 1.**
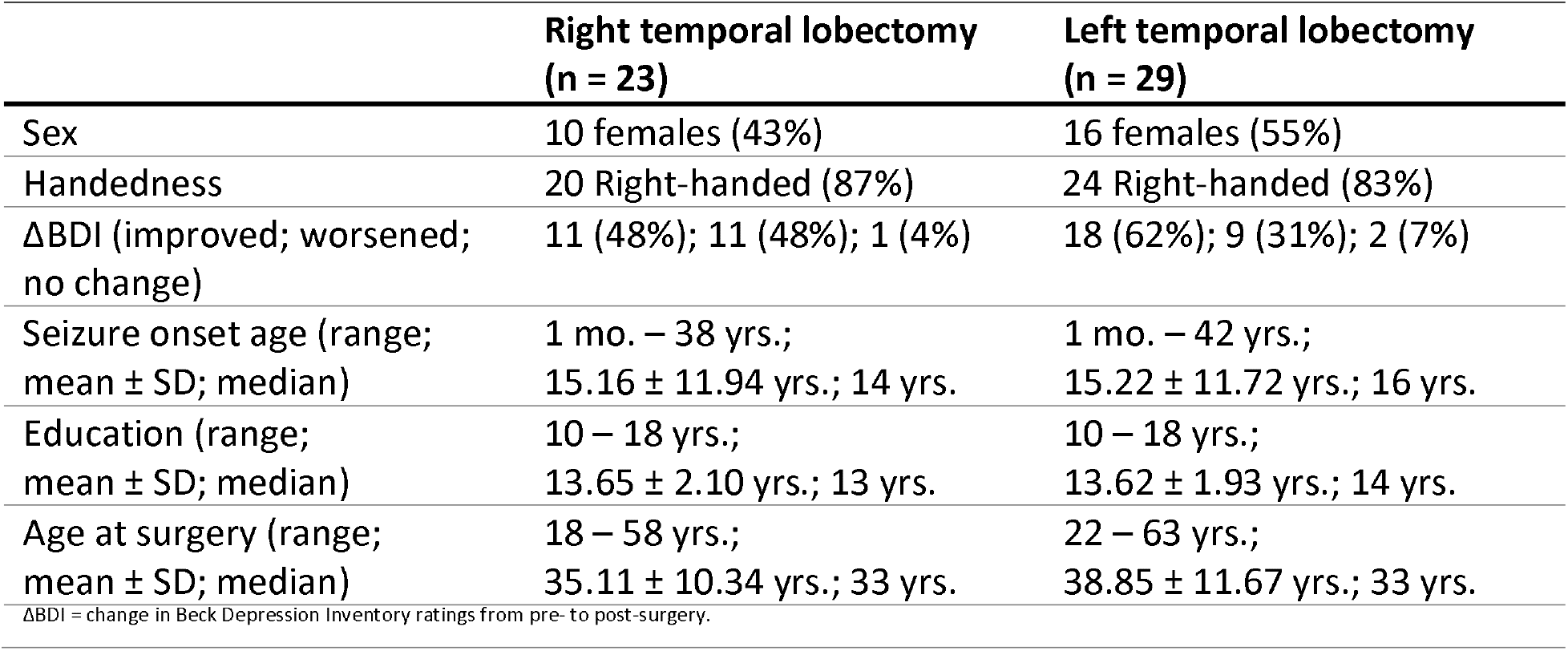
Description of participants who underwent a successful medial temporal lobe resection and achieved post-surgical seizure freedom during the first year.

### Lesion segmentation

All participants had a medial temporal lobe resection with visible lesion boundaries evident from a research-quality structural MRI (T1 and T2 sequences). The surgical resection cavity was traced manually for each participant using the MAP-3 method of lesion tracing for individuals who enrolled in the Patient Registry prior to 2006, which involves manual tracing of lesion borders on a template brain (Damasio and Frank, 1992; Fiez *et al*., 2000). After 2006, lesions were manually traced on native T1-weighted scans (Smith *et al*., 2004) and then transformed to the 1-mm Montreal Neurological Institute (MNI-152) template brain (fsl.fmrib.ox.ac.uk/fsl/) using nonlinear registration and lesion masking techniques available in ANTs (Avants *et al*., 2008). The anatomical accuracy of the lesion boundaries was reviewed and edited as needed in native and MNI space by a neurologist (A.D.B.) blinded to mood rating outcomes.

### Lesion analyses

A data-driven multivariate lesion symptom mapping of mood change, measured as the difference in BDI scores from pre- to post-surgery, was used to identify brain structures that, when lesioned, are significantly associated with changes in BDI. We used the LESYMAP package in R (https://github.com/dorianps/LESYMAP), which uses a sparse canonical correlation analysis for neuroimaging (SCCAN) (Pustina *et al*., 2018). The SCCAN method involves an optimization procedure to assign a continuous weight to each voxel (range 0 – 1) in order to maximize the multivariate correlation with the true behavioral scores. The validity of the map is evaluated using a 4-fold cross-validation (with 25% of the sample is held out at each fold). The optimal sparseness and statistical significance of the lesion-behavior association is tested by comparing the actual change in mood to that predicted by the model. Significance is tested on the entire map, which avoids problems associated with multiple comparison correction. This method has been previously validated and demonstrated to be more spatially precise relative to a mass univariate approach (Pustina *et al*., 2018). The multivariate analysis was conducted with all participants combined and for each hemisphere run independently to account for potential differences related to lesion laterality. We also generated a proportional subtraction map to show regional changes in the lesion locations associated with changes in BDI ratings for descriptive purposes. The lesion overlap map of individuals with improved mood was subtracted from the lesion overlap of individuals with worsened mood. The voxels remaining from the subtraction were damaged at a proportionally higher rate in participants with mood improvement (e.g. if a voxel was affected in 70% of improved individuals and 5% of worsened individuals, then this voxel would have a difference value of 70% − 5% = 65%). This qualitative analysis can display regional trends that may not reach statistical significance in the multivariate lesion-symptom mapping analysis.

## Results

Overall, participants showed a significant post-surgical improvement in mood with lower post-surgical BDI ratings (mean ± SD: 8.37 ± 7.38 points, median 6 points) compared to pre-surgical ratings (mean ± SD: 11.75 ± 9.53 points, median 9 points); paired sample t-test (p = 0.045). Post-surgical mood improvement was observed in 56% of participants (n = 29; mean ± SD: 11.28 ± 8.99 points, median 9 points), mood worsening in 38% (n = 20; mean ± SD of 7.55 ± 6 points, median 5.5 points), and no change in mood assessment in 6% (n = 3). An independent sample t-test and a chi-square test showed no significant relationship between the ΔBDI with lesion laterality, handedness, or gender (p > 0.05).

### Lesion analyses

The location of the brain lesions from the 52 individuals included in the analysis is shown in Fig. 1a. The lesion symptom mapping analysis of right medial temporal lobectomy lesions was the only analysis that reached a statistical significance (r = 0.5, p = 0.008). The map highlighted the right amygdala as the region most strongly associated with mood improvement, including the ventral basolateral nucleus and periamygdaloid cortical transition area (Tyszka and Pauli, 2016), with a peak voxel at MNI coordinate (15, −5, −21), Fig. 1b. The proportional subtraction analysis was similar in highlighting the same region of the right amygdala with peak coordinates at (14, − 5, −23), Fig. 1c. The same pattern of association was also seen on the left hemisphere with a left amygdala peak at (−22, −5, −19), but this did not reach a statistical significance in the LESYMAP analysis. Additional findings that were observed but were not a part of our *a priori* hypothesis included a significant association of lesions of the right hippocampus and surrounding cortex with higher post-surgical BDI ratings, including perirhinal ectorhinal cortex (22, 0, −35) and the parahippocampal area 1 (21, −38, −10) (Glasser *et al*., 2016).

**Fig. 1.**
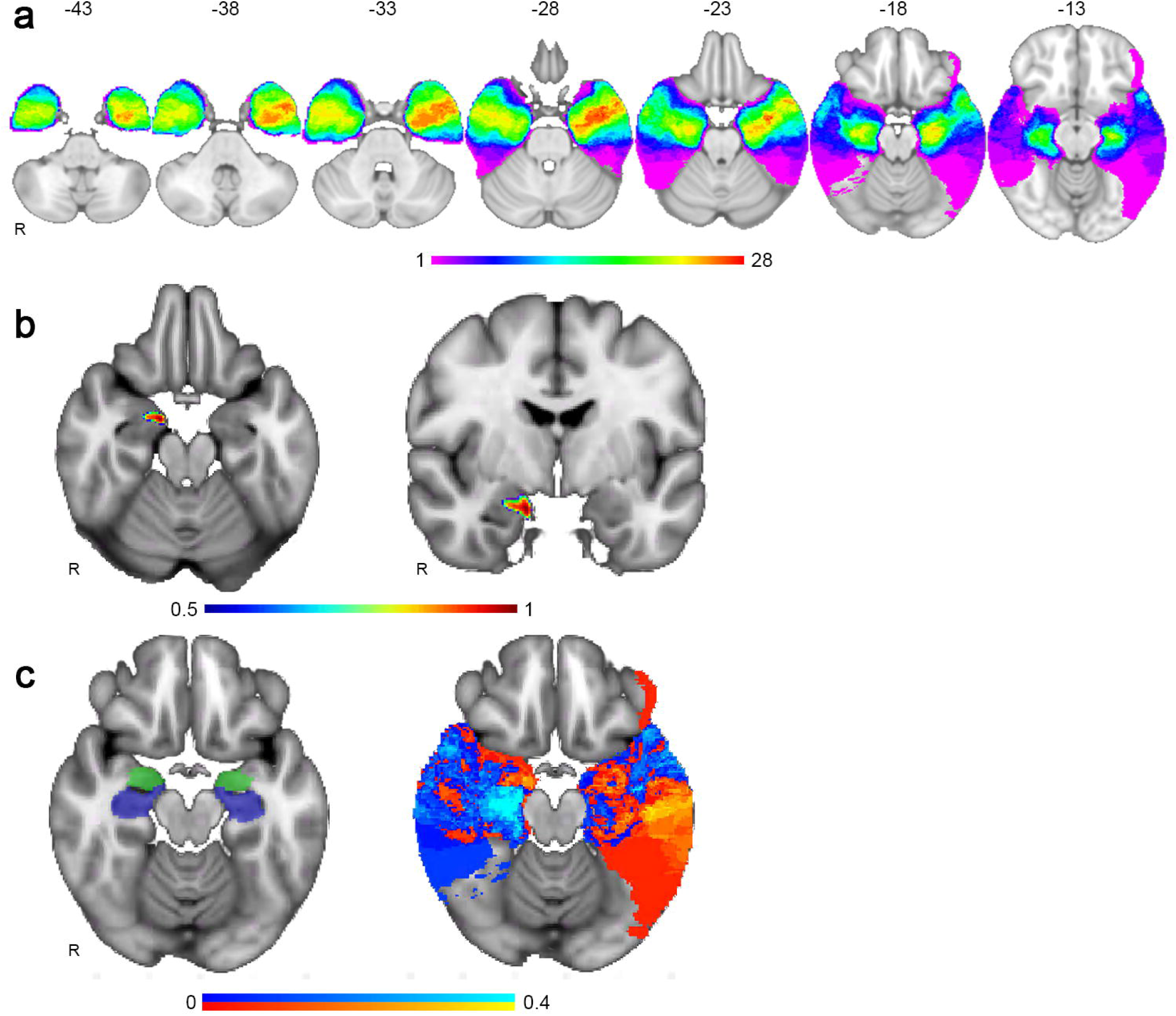
**a. Lesion overlap.** Lesion overlap map of the surgical resection cavities used in this analysis (N = 52), with a peak overlap in the left medial temporal lobe n = 28 of 52 at (−40, −4, − 25). **b. Multivariate lesion symptom mapping**. The lesion symptom map of participants with right hemisphere lesions, showing an association of right amygdala lesions with improvement in post-surgical mood, with voxel weights displayed using a unitless scale with values closer to 1 reflecting a stronger lesion-mood association. (r = 0.5, p = 0.008). **c. Lesion proportional subtraction map**. The image on the left shows the location of the amygdala and hippocampus in green and blue, respectively, as a reference. The image on the right shows proportional subtraction results, where the red color scale displays regions preferentially associated with improved mood and the blue color scale displaying regions associated with mood worsening. Note the sharp contrast that respects that anatomical boundary between the hippocampus and amygdala, with hippocampus lesions more commonly associated with mood worsening and amygdala lesions associated with a higher likelihood of post-surgical improvement in mood.

## Discussion

This study examined lesion location as it relates to changes in mood in patients who underwent medial temporal lobectomy for epilepsy. We hypothesized that amygdala lesions would be associated with improvement in self-reported mood, as assessed with BDI ratings. Our hypothesis was supported for the right, but not the left, amygdala, as lesions of the right amygdala were significantly associated with improvement in mood based on multivariate lesion symptom mapping. These findings are consistent with the role of the amygdala in processing negative emotions and psychopathology, including symptoms of depression (Donegan *et al*., 2003; Tranel *et al*., 2007; Victor *et al*., 2010). This builds on prior studies of mood outcomes following a medial temporal lobe resection (Spencer *et al*., 2003; MacRodimitris *et al*., 2011; Smith *et al*., 2018; Bijanki *et al*., 2020; Hebel *et al*., 2021). To the best of our knowledge, this is the first study to investigate the effects of surgical resection of the amygdala on mood using a multivariate lesion symptom mapping method, which provides a data-driven approach for relating change in mood symptoms to specific medial temporal lobe structures that, when resected, are significantly associated with these changes.

Our findings also suggest that surgical resection of the right hippocampus and adjacent medial temporal lobe cortex was associated with increased depression symptoms, which also has precedent in the literature (Yang *et al*., 2007; Cheng *et al*., 2018; Rolls *et al*., 2020). Taken together, these findings support the notion that medial temporal lobe structures have a causal role in mood, with lesions of the amygdala and hippocampus/parahippocampal gyrus potentially having opposing effects on mood. This is supported by the sharp contrast at the amygdalo-hippocampal border that shows an association with mood improvement and worsening, respectively (Fig. 1c). A limitation in the analysis is a lack of cognitive outcome data, so the influence of cognitive changes on mood is unclear. Further prospective studies of large cohorts will be useful in further characterizing the role of medial temporal lesion location and mood outcomes.

## Data Availability

The datasets generated and/or analyzed during the current study are available upon request from the corresponding author.

## Acknowledgements

The authors thank the patients who participated in this research and thank Dr. Ralph Adolphs for his scientific feedback.

## Author Contributions

F.M.A. conducted the data analysis and drafted the manuscript and figures. D.T. and M.A.H. contributed to the data acquisition and the revision of the manuscript. A.D.B. contributed to the conceptualization, design, and supervision of the study. All authors revised and approved the final version of the manuscript.

## Ethics approval

All procedures performed in the study were in accordance with the University of Iowa Institutional Review Board.

## Consent to participate

Informed consents were obtained from all participants in accordance with the University of Iowa Institutional Review Board.

## Consent to publish

Not applicable.

